# Large-scale analysis of the medical discourse on rheumatoid arthritis: complementing a socio-anthropologic analysis

**DOI:** 10.1101/2024.07.02.24309823

**Authors:** Mario Santoro, Christine Nardini

## Abstract

The medical discourse, entails the analysis of the modalities, far from unbiased, by which hypotheses and results are laid out in the dissemination of findings in scientific publications, giving different emphases on the background, relevance, robustness, and assumptions that the audience should take for granted. While this concept is extensively studied in socio-anthropology, it remains generally overlooked within the scientific community conducting the research. Yet, analyzing the discourse is crucial for several reasons: to frame policies that take into account an appropriately large screen of medical opportunities, to avoid overseeing promising but less walked paths, to grasp different types of representations of diseases, therapies, patients, and other stakeholders, understanding and being aware of how these very terms are conditioned by time, culture and so on. While socio-anthropologists traditionally use manual curation methods, automated approaches like topic modeling offer a complementary way to explore the vast and ever-growing body of medical literature. In this work, we propose a complementary analysis of the medical discourse regarding the therapies offered for rheumatoid arthritis using topic modeling and large language model-based emotion and sentiment analysis.

## 1. Introduction

The medical discourse [1,2] represents an aspect of scientific communication somewhat neglected by the very community producing the research. In particular, it entails an analysis of the modalities by which hypotheses and results are presented in the communication and dissemination of findings, adding more or less conscious bias. Exploration of the discourse is important for decision-making based upon such scientific evidence and hence for policy making, for funding orientation and so forth, to know better how these terms are conditioned by time, culture and so on.

We here propose the analysis of the medical discourse on therapies for the treatment of rheumatoid arthritis (RA), as an exemplar inflammatory, autoimmune, and hence non-communicable disease (NCDs). NCDs are among the most deadly and burdensome maladies affecting transversally and worldwide societies [3]. For this reason, their control is among the objectives of the WHO Sustainable Development Goal (SDG) 3.4. To perform our analysis, we explore a spectrum of clinical approaches including three large categories: pharmacological (PHA) and non-pharmacological (including physical, generally unstandardized -USTD- and experimental, EXP) RA therapies. In short, in addition to the therapies directly addressing the immune system response (innate, adaptive and trained), we include additional modulators, i.e., the autonomic nervous system (ANS), the gut intestinal (GI) microbiome and the elicitation of the wound healing processes as proposed in [4]. Based on this general framework, PHA approaches to tackle RA represent by far the most assessed and recognized category, i.e., the gold standard in the most widespread practice supported by scientific societies in the United States of America (American College of Rheumatology, ACR), Europe (European League for Rheumatoid Arthritis, EULAR) and Asia-Pacific (APLAR). PHA targets the immune system response and includes a large set of drugs controlling inflammatory symptoms, with the following sub-categories: non-steroidal anti-inflammatory drugs (NSAIDs), paracetamol or morphine (-derived) analgesics; corticosteroids to counteract the degeneracy of the disease; disease-modifying anti-rheumatic drugs (DMARDs) to interfere or block the pro-inflammatory endogenous activity [5–7] in a generic (conventional cDMARDs) or targeted (biologic, bDMARDs) way. Experimental (EXP) therapies are mostly concerned with the attempts to control inflammation via interactions with the ANS(vagus nerve stimulation -VNS) and the GI microbiome (diet, nutraceutics, antibiotics, and fecal microbial transplant -FMT, and, initially, antibiotics, by now dismissed). Finally, wound healing modulators are represented by physical (optic, mechanic, magnetic, and electric) stimuli whose therapeutic release, although extremely promising under certain medical specializations (see, for instance, Alzheimer’s Disease [8,9] suffers to date from limited standardization, evidence-based research, guidelines, funding, and overall public recognition (unstandardized category -USTD). Our current work builds on a recent effort [10] where we performed the analysis of the medical discourse based on the manual curation of two exemplar articles per subcategory of therapy for a total of 28 articles. In that work, our conclusions highlighted some interesting non-trivial patterns, including, for instance, the lack of awareness among USTD practitioners of the basic biology scientific evidence underlying their therapies, or the bias in meta-analyses approaching the same therapies (electrostimulation) depending on the traditional/modern medical perspective adopted, to name few remarkable examples. However, one of the main limitations of that study lies in the reduced amount of literature analyzed (for details, see Materials and Tables S1-2).

To overcome this, topic (Vayansky and Kumar, 2020) and large language models (LLM, [11]) represent effective approaches to perform textual analyses on a large amount of data. Therefore, we present an application of these approaches to explore the totality of the material selected in [10], i.e. 204 articles (from now on the corpus, see Materials and Methods and Table S3). Briefly, we employ Structural Topic Modeling (STM,) (Roberts et al., 2013) to automatically uncover latent thematic structures within the corpus. STM offers a more sophisticated approach compared to simple keywords identification. By analyzing co-occurrence patterns of words across documents, STM reveals underlying thematic relationships that may not be readily apparent through manual reading.

Furthermore, we incorporate Large Language Models (LLMs) to enrich the analysis. LLMs, such as those available through the Hugging Face Transformers library [12], are trained on massive text datasets, enabling them to perform various natural language processing tasks with exceptional accuracy. In this study, we leverage the capabilities of LLMs to perform three main tasks: language recognition, translation, and emotion analysis.

To perform this analysis, we developed an original pipeline using Semanticase http://www.semanticase.it PiazzaCopernico 2023 (developed as a commercial tool by author MS, and whose methods’ implementation has not been discussed elsewhere). Semanticase first automatically identifies the topics emerging from the literature corpus. The lexical semantics (EXP, PHA, USTD) are analyzed category-wise with the computation o fseveral statistics c, and finally the topics sentiments’ and emotions’ content are explored via text-based and LLM models, respectively. In the process new functionalities have been developed, now added to Semanticase (see Methods).

Overall, this work returns an analysis that complements and enriches the previous one regarding the categories’ characterization. In particular, the computational approach was able to successfully identify several subcategories not given a priori (i.e. not given as covariates, one example for all conventional and biologic DMARDs). Also, the approach was able, conversely, to merge subcategories that practice keeps separate (namely, electroacupuncture and VNS). Finally, the newly run emotions analysis was able to discover a similarity between PHA and EXP and versus USTD that we could relate to the temporal legitimation of the manual approach, suggesting that the medical discourse is strongly biased by these variables, relating to the approval therapies receive, based on the features that are perceived to give them strength (long dated practice versus exploitation of state-of-the art innovation).

## 2. Materials and Methods

### 2.1. Materials

The starting point to identify the materials was the one described in [10]. Briefly, the selection emerged from a literature search on PubMed performed by three-tiers queries using Medical Subject Headings (MeSH [13]). The first tier is a general query on “Arthritis, Rheumatoid/therapy”[MeSH] AND “Review” [Publication Type]”. The latter tiers (Level I and II) specialize the former with specifiers that include (level I) the mechanism modulating inflammation [4] and their physical nature (level II). Level I includes the GI microbiome, the ANS and wound healing. Level II is taken by the list of stimuli (optic, mechanical, electric, magnetic) routinely used in wound healing testing [14].

Since our ambition was to use all the corpus (500 articles along with their PubMed Identifiers, PMIDs), and not a representative selection (28 in [10]), in this work we have further processed the list and cleaned it from 31 duplicates leading to 469 articles. Further, to retain only research articles (and avoid other types of publications) only 347 articles with abstracts were retained. Finally, due to varying levels of accessibility, only 204 articles (58.8% of the originally identified PMIDs) could be fully downloaded for further analysis. As covariates in the dataset, we used the three EXP, USTD, and PHA categories, thus dividing the articles into 3 groups. The original corpus was converted into plain text from its original PDF format using Semanticase. The original list of 500 articles along with the Pubmed queries and all categories and subcategories can be found in Table S1, the list of the 28 manually curated articles in Table S2, the list of the final 204 PMID analyzed in this article in Table S3.

### 2.2. Methods

#### 2.2.1. Sentence Splitting

To facilitate a more granular text handling and analysis, we developed a sentence-level approach. This strategy allows for applying various techniques specifically designed for sentence-based data. Pysbd, a well-established Python library renowned for its proficiency in sentence boundary detection, is the cornerstone of this process. By utilizing pysbd’s functionalities, we achieve accurate document segmentation, resulting in well-defined sentence units that form the basis for subsequent analyses. This novel approach has been now (contextually to this work) added to Semanticase software.

#### 2.2.2. Corpus Preprocessing

Corpus creation for a domain like analysis of scientific papers often necessitates an iterative approach to refine the data and identify potential inconsistencies progressively to safeguards both comprehensiveness and accuracy. This includes the management of: (i) multilingual articles; (ii) frequent but non meaningful acronyms or other types of symbols; (iii) synonyms that inflate the statistics on words’ frequency and from there the inference of semantics.

Regarding multilingual articles, the sentence-level strategy is crucial to identify all and only sentences that are not in English. Subsequently, we leverage Semanticase’s language identification capabilities, to identify documents containing non-English sentences that are then translated using Semanticase itself, and finally reassembled in the original order to reconstruct the entire document in English. Although this has highly enhanced the quality of the translation, results are not perfect, nevertheless, topic modeling (see Results) was able to group untranslated sentences within one topic only (Topic 3).

Regarding acronyms/symbols and synonyms the corpus undergoes a rigorous preprocessing to optimize data quality, beginning with text normalization, ensuring consistency in word representation by converting all text to lowercase. Subsequently, a regular expression-based cleaning routine meticulously eliminates extraneous characters and symbols that could potentially introduce noise during analysis. Finally, to enhance domain-specific comprehension, a human-in-the-loop synonym identification and substitution process is implemented. Similarly, meaningless symbols and acronyms are manually removed. This iterative curation, informed by basic word analyses, aims to standardize the corpus vocabulary and ensure that the nuanced terminology of the specific domain is effectively captured.

#### 2.2.3. Words Statistics

We employed a statistical technique known as keyness analysis [15] to explore the distribution of words across the corpus and identify terms potentially characteristic of specific document categories. Keyness analysis quantifies the differential occurrence of words between a target group and a reference group within a corpus. In this context, the Semanticase web application leverages the textstat_keyness function from the quanteda.textstats R package [16]. This function further calculates a chi-squared statistic to assess the significance of the observed difference in word frequency between the target and reference groups.

Yates’ correction is applied to the chi-squared statistic to mitigate potential biases introduced by small sample sizes within individual documents. For a more in-depth exploration of keyness analysis and its applications in text analysis, we refer readers to [17].

This study’s target group corresponds to the category (i.e., PHA, EXP, USTD).

Conversely, the reference group encompassed the combined word frequencies across all other documents in the corpus. This approach aims to identify words that exhibit statistically significant higher frequency within a particular category than the overall corpus, potentially reflecting the thematic focus of that category.

#### 2.2.4. Topic Modelling

Our study employs topic modeling to uncover latent thematic structures within the corpus. For a comprehensive overview of the Structural Topic Model (STM), we refer to Roberts et al. (2014).

As above, we leverage a specific covariate, i.e. the category (EXP, PHA, USTD), within the STM framework. This covariate allows us to quantitatively estimate the emerging thematic structures (a.k.a. topics) associated with different article types within the corpus. Incorporating this covariate gives us more profound insights into how thematic structures vary across different article categories. In particular, we used two defined statistics to quantify this insight.

The first, is the topic prevalence and refers to the degree to which a particular topic is represented within a document or across the entire corpus. It measures how frequently or prominently a topic appears in a text, and using STM, we measure topic by topic whether there is a stronger or weaker association with a specific category.

The second is the topic content that encompasses the specific N-grams (set of N consecutive words, with N=1,2,3) that define a particular topic. It represents the vocabulary that characterizes a concept or a theme. STM defines the probability for each word in the topic to be associated to each covariate (category). Importantly, this enables the possibility that few of the words in a topic are more (high probability) or less (low probability) associated to a specific category. This indicates that, although the topic is important across the whole corpus, i.e. shared among all categories, different words or N-grams may be characteristic and descriptive of this topic, category-wise. I.e. a corpus topic can be described by a partially different vocabulary, category-wise.

The algorithm automatically identifies the number of topics in Semanticase, provided a range is given. Here also, human-in-the-loop served to identify the most suitable range, which was set to [10-19].

Following the execution of the topic model, Semanticase exploits the STM inter-topic correlations (directly output from the model) as measures of similarity among the topics and constructs a hierarchical clustering tree. Using an aggregative logic, each leaf is associated with the most probable words that are shared between the topics’ group, recursively.

#### 2.2.5. Sentiment Landscape through Sentence-Level Analysis

We employ the sentence spitting defined above and the sentiment analysis framework within the Semanticase software to enrich the emotional analysis, leveraging the sentimentr R package [18]. Lexicon-based sentiment analysis relies on pre-defined sentiment lexicons, categorizing words and phrases into positive, negative, or neutral. It is crucial to acknowledge the inherent limitations of this approach: sentiment analysis can be nuanced, and the presence of valence shifters – such as negation words (“not,” “never”), intensifiers (“very,” “extremely”), and adversative conjunctions (“but,” “however”) – can significantly alter the intended sentiment within a sentence. The sentimentr R package incorporates rules to mitigate the influence of some valence shifters. This lexicon-based analysis provides a valuable starting point for understanding the broader emotional trends within the corpus (see below).

#### 2.2.6. Emotional Landscape through Sentence-Level Analysis

We exploit the emotion analysis (more nuanced than the three-tiers sentiment analysis) to better understand the feelings landscape within the corpus, at sentence level. We employ for this a pre-trained classification pipeline built upon the huggingface transformer [19] roberta-base-go_emotions model [20]. This model, leveraging the comprehensive GO Emotions dataset [21], encompasses 27 distinct emotional states plus a neutral category (Table S4).

The employed model outputs probability scores (between 0 and 1) for all 27+1 emotions. In specific scenarios, a sentence might lack a dominant emotional presence, i.e., exhibiting scores like 0.5 and 0.49 for two emotions, with the remaining scores being significantly lower (<0.01). Capturing the nuance of such co-occurring emotions might be valuable for a comprehensive analysis. Recognizing the inherent polysemy of individual sentences, where multiple emotions may be conveyed with varying intensities, we establish a threshold of 0.3, posing that a maximum of three concurrent emotions can be incorporated into the analysis, provided their respective scores surpass the threshold of 0.3. This threshold serves as a selectivity mechanism, retaining only emotions exceeding this probability for further analysis. This approach aligns with the well-founded assumption that a single sentence is unlikely to harbor many strong emotions concurrently.

A normalized sentence index (sent) is devised to facilitate comparative analysis across papers’ categories. This index assigns a unique, sequential integer to each sentence within a paper. Subsequently, this value is normalized by dividing it by the total number of sentences in that paper. This approach yields a sent value ranging from 0 to 1 for each paper.

Finally, to visualize for each emotion, the category trends (article by article, probability scores across sentences, progressively), we utilize ggplot2 [22]. We generate smoothed line graphs with default parameters of the geom_smooth function, a local regression smoother (loess), which fits a low-degree polynomial to subsets of nearby points, to enhance plot readability.

## 3. Results and Discussion

### 3.1. Topic Modeling

The analysis automatically returns 10 topics, whose hierarchical shape appears to be almost fully nested (with the exception of Topic 1 and 8), as it is shown in Figure 1. The structure is represented according to a hierarchical cluster, and shows that every topic is fully included in the topic above (Topic 3 included in Topic 2, included in Topic 10 etc.). This possibly indicates that the structure identified proceeds by progressive specialization of one major topic (Topic 6), rather than by the identification of a certain number of topics with similar relevance, which would be represented by more than one topic at the same height (as it is the case for Topics 1 and 8).

**Figure 1.**
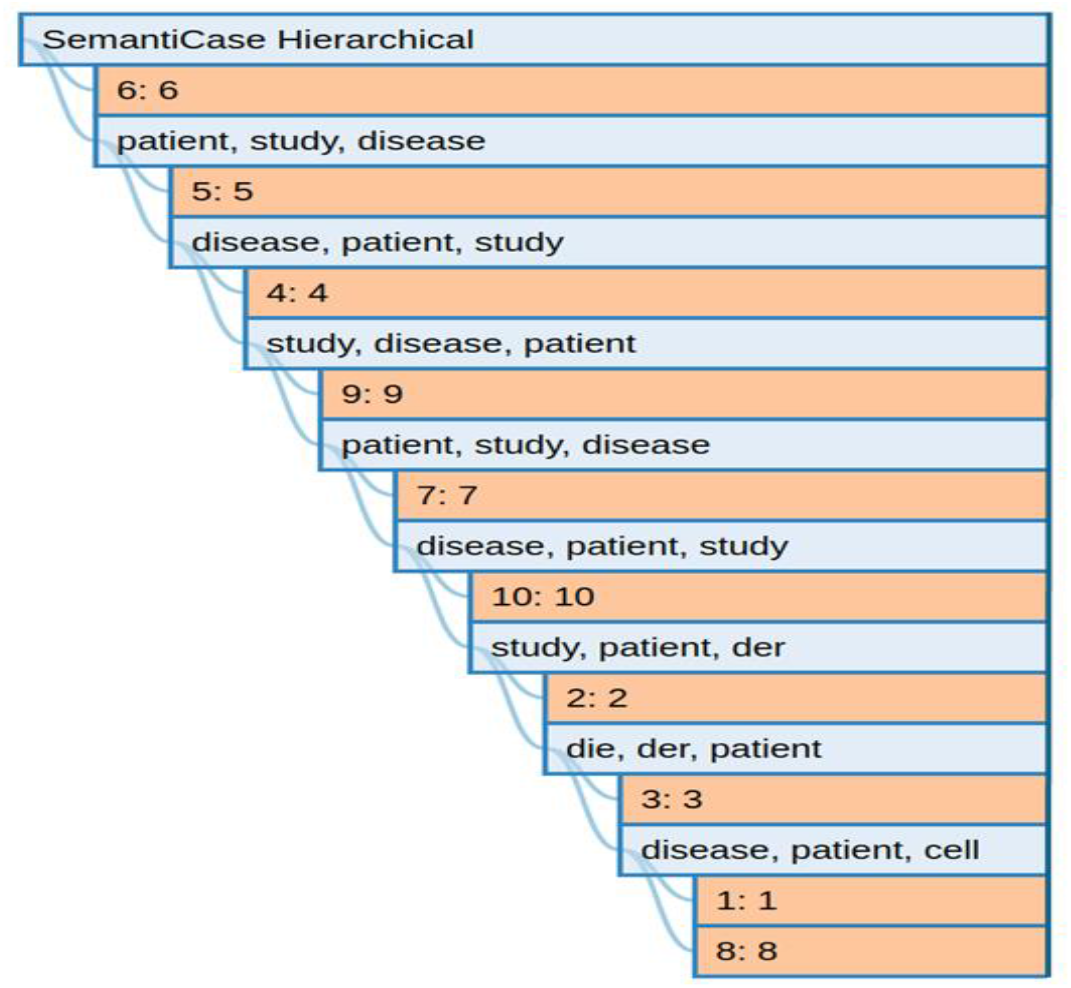
Topics hierarchical clustering. Construction starts from Topics 8 and 1 (highest correlation) and reports the words shared by the two topics with highest probability (disease, patient, cell).

To ease the discussion, we present an extract from within the topics’ words statistics to enable an intuitive Topic labeling (label column, Table 1). The selection in the Words column is custom-made, based on a relatively short list of words (from 10 to 15 per topic), guided by the meaningfulness of the words themselves. Other labeling is clearly possible, but we figured this naming is an appropriate compromise for readability. For this reason the variables LIFT (high frequency within the topic) and score (exclusivity to the topic) have been discarded owing to: the “dirty” selection (acronyms, numbers, left over despite the preprocessing, see Methods, whose interpretation is irrelevant or non-intuitive), for the former, and redundancy with Prob and FREX for the latter. We nevertheless encourage the reader to explore the full set of results in Table S5.

**Table 1.**
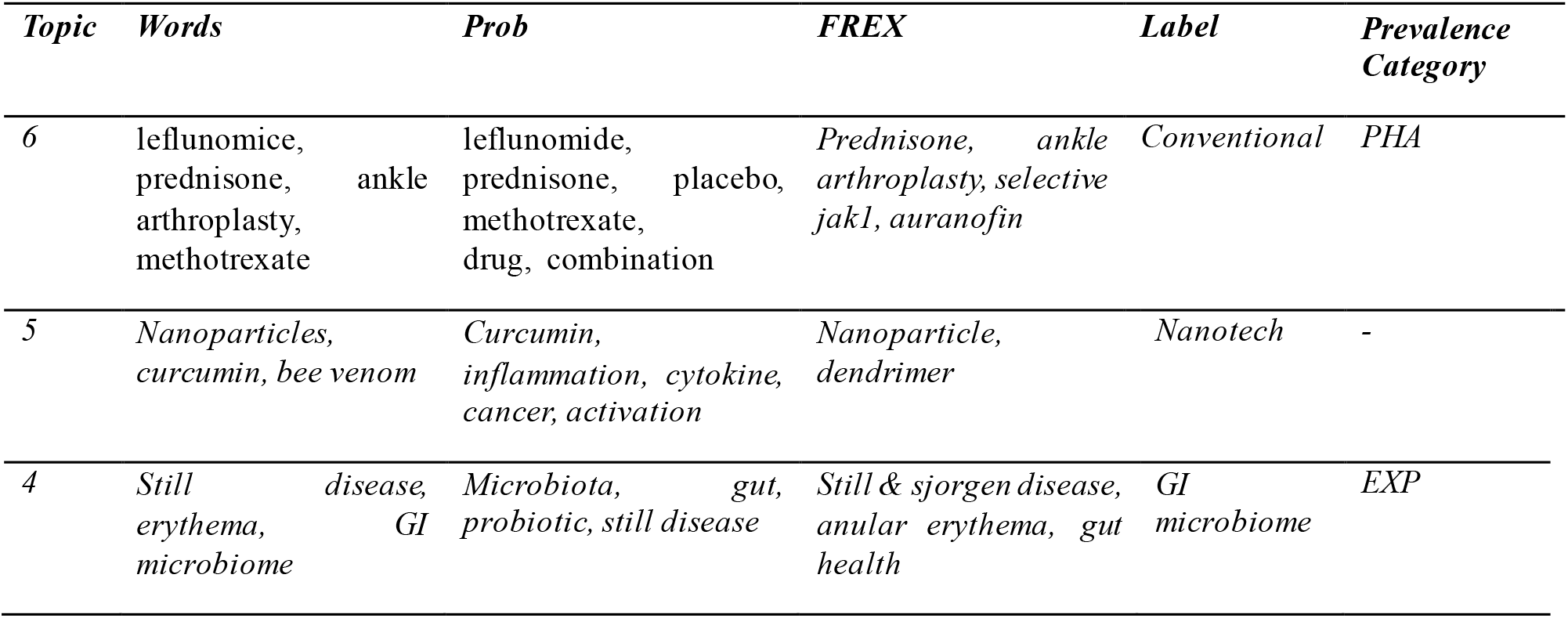
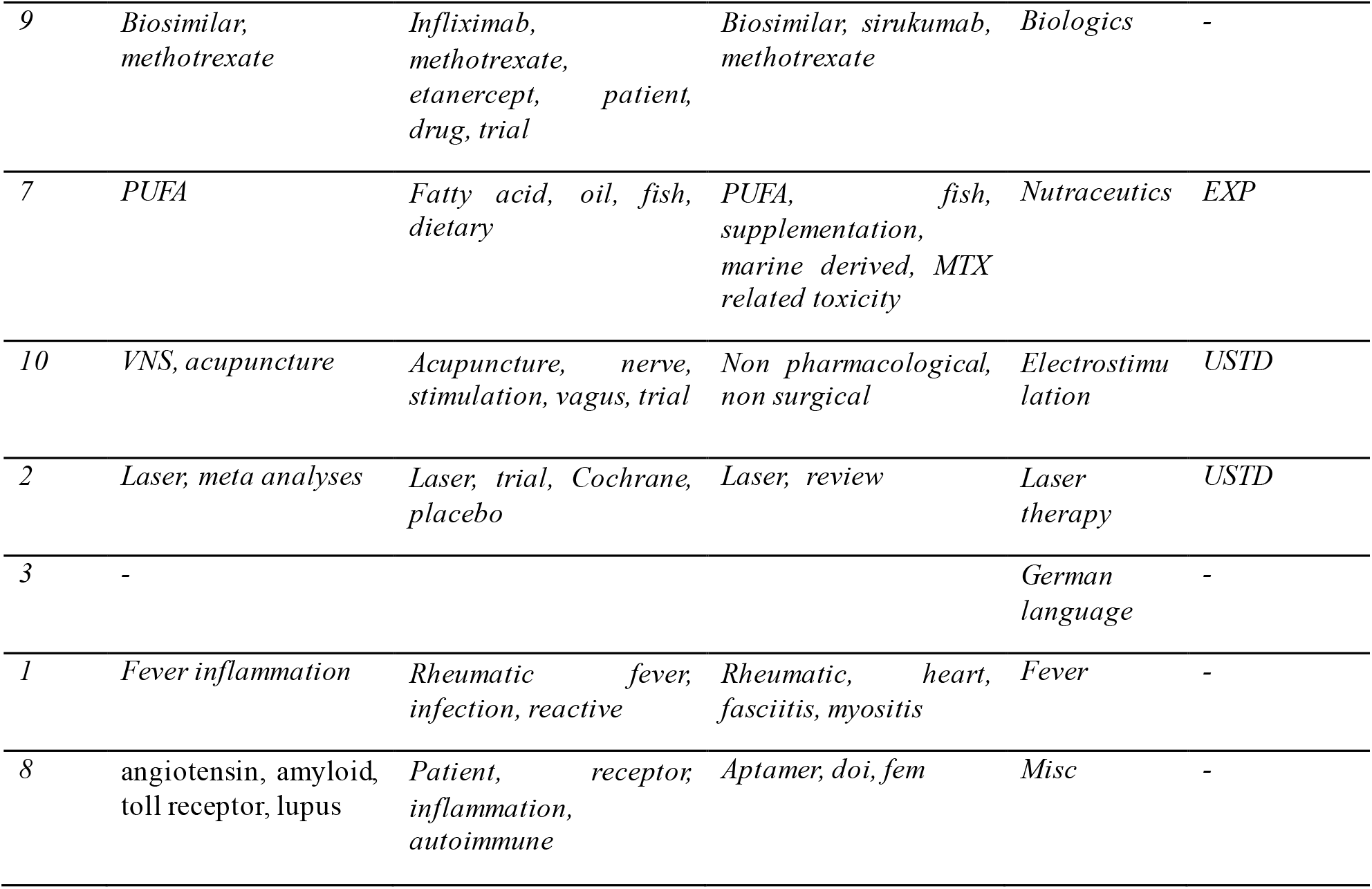
Topics labels and main statistics.

This hierarchical structure is in line with the observed dominance of the mainstream clinical approach to RA (overlapping with category PHA, synonym of conventional). This approach, as well as the associated scientific narrative, includes a common and widespread model to present the Introduction to virtually all articles, represented by the setting of the problem with a conventional clinical definition and etiology of the disease. Within this structure (Fig 1) other approaches/therapies appear to be nested, with a language possibly reporting on innovation (EXP) including in Topic 5 (nanomedicine, as well as -on lower ranks-VNS (expectedly) and curcumin (expected in the nested Topic 7)), then GI associated therapies (Topic 4) and biologics (Topic 9, includes in addition to the cDMARD methotrexate also bDMARDs, representing the innovation within conventional medicine) and finally nutraceutics (Topic 7). From this point on, non-pharmacological/physical therapies are identified including acupuncture & VNS(Topic 10) and laser (Topic 2). Follows cluster/Topic 3 with German written articles where automatic translation fails, which includes Topics 1 and 8, more difficult to label, relatively heterogeneous and less prevalent (see Expected Proportion in Table 2).

**Table 2.**
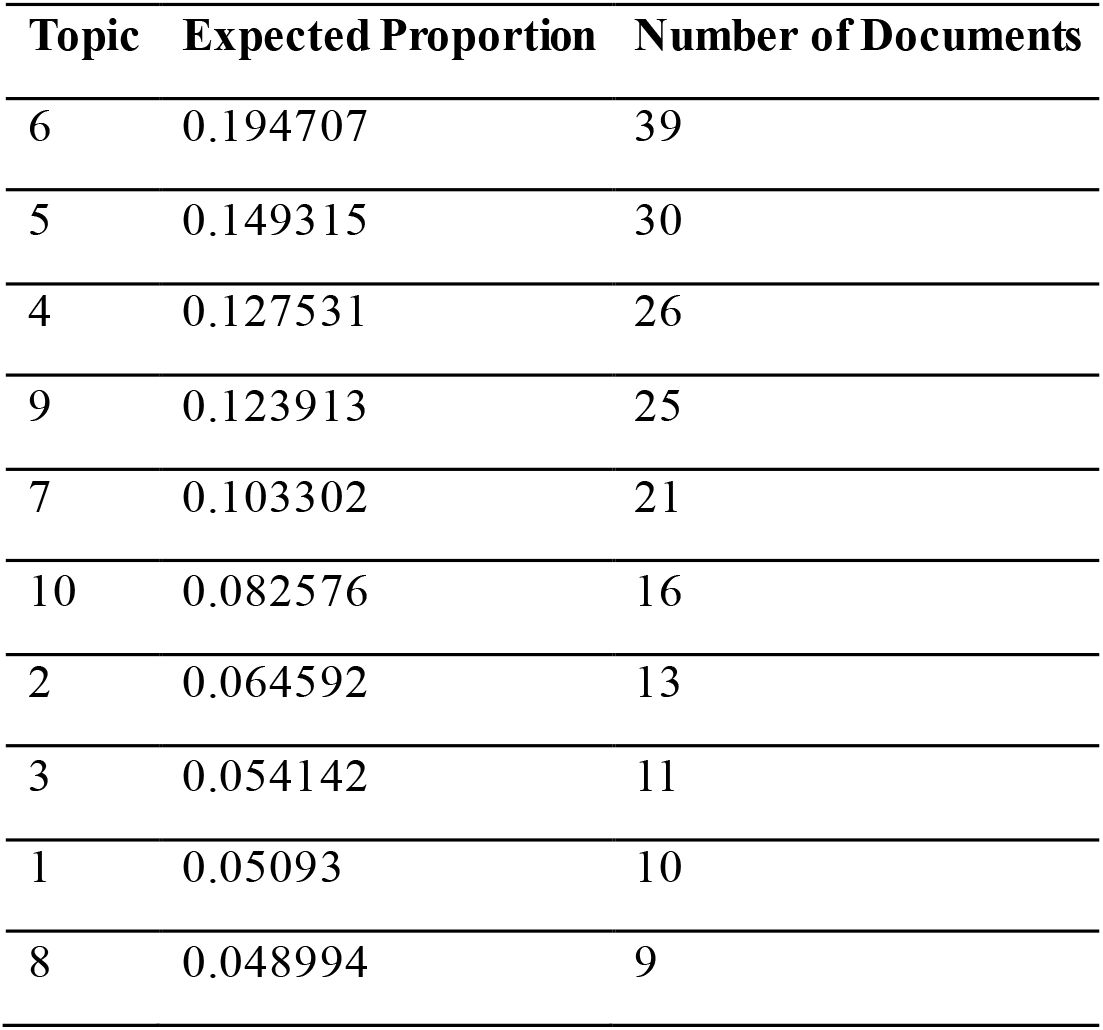
Topics proportion, by number of documents in the corpus.

| Topic | Expected Proportion | Number of Documents |
| --- | --- | --- |
| 6 | 0.194707 | 39 |
| 5 | 0.149315 | 30 |
| 4 | 0.127531 | 26 |
| 9 | 0.123913 | 25 |
| 7 | 0.103302 | 21 |
| 10 | 0.082576 | 16 |
| 2 | 0.064592 | 13 |
| 3 | 0.054142 | 11 |
| 1 | 0.05093 | 10 |
| 8 | 0.048994 | 9 |

Observations on the prevalence of such topics (Table 1, col 4) confirm once more that the above hierarchy (conventional, innovative, non-pharmacologic) is mirrored within categories, with a dominance of PHA over EXP over USTD, and topic proportion (Table 2), where we observe that Topic 6 dominates (39 documents), followed by specialization in Topics 5, 4, 9, 7 concerned mainly with innovation with 30, 26, 25 and 21 documents respectively, followed by Topic 10 (16 documents) including acupuncture and VNS, and Topic 2 (13 document, laser), finally followed by Topics 3, 1, 8 (11, 10, 9 documents, respectively).

When compared to our previous work [10] we can observe that several of the subcategories we defined a priori (reported in Table 3 for convenience) have indeed been automatically identified. In particular: Dys (dysbiosis, Topic 4) and Diet (Topic 7). Further, the approach was able to discriminate between conventional and biologic DMARDs, (Topics 6 and 9 respectively, which were merged under the AI label). Interestingly VNS, EL and AP are all merged under Topic 10. Additional topics (that were not discussed in [10]) are concerned with nanotechnology (Topic 5) and fever (Topic 1).

**Table 3.** Listing of subcategories identified manually in [10].

| Category | Label | Extended MeSH |
| --- | --- | --- |
| PHA | AI | Anti-inflammatory Drug Therapy |
| EXP | VNS | Vagus Nerve Stimulation |
| EXP | Dys | Dysbiosis Therapy |
| EXP | AB | Anti-Bacterial Agents |
| EXP | Diet | Dietary Supplements |
| EXP | FMT | Fecal Microbiota Transplantation |
| USTD | US | Ultrasonic Therapy |
| USTD | Mass | Massage |
| USTD | AP | Acupuncture Therapy |
| USTD | EL | Electric stimulation Therapy |
| USTD | LLT | Low Laser Therapy |
| USTD | EM | Electromagnetic Phenomena |

Finally, AB (antibiotics), FMT (fecal transplant), US (ultrasound), MASS (massage), EM (electromagnetism) did not emerge, with the exception of AB, this can most likely be attributed to the very limited amount of literature available (they were indeed supported by 121, 0, 5, 2, 1 articles, respectively, in our original work), and AB being a dismissed therapy and hence discussed in old articles only a limited number (42) was included among the 204 we could work with.

### 3.2. Sentiment Analysis

Regarding sentiments (Fig. 2), PHA presents with a relatively large plateau on the slightly negative sentiment side, EXP is definitely negative and USTD presents a slightly bimodal positive behavior.

**Figure 2.**
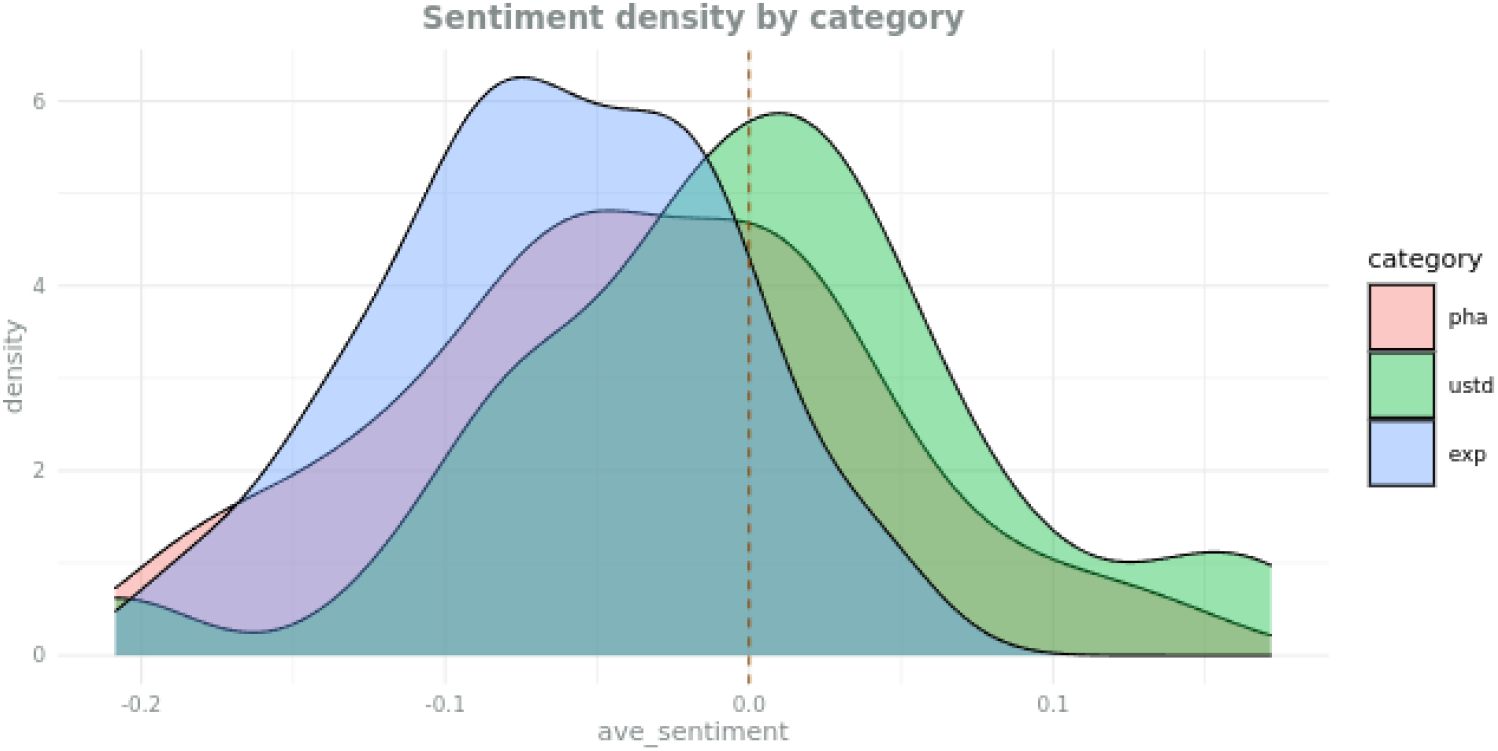
Sentiment distribution across the literature corpus, by categories

When observing how sentiment are distributed by topic (Table 4), upon removal of the uninformative (German language) Topic 3, we observe that the most negative average sentiment are shared by fever and nutraceutics (Topics 1 and 7) followed by comparably milder negative sentiment in all other Topics, with the exception of conventional and (nerve) electro-stimulation, (Topics 6 and 10), characterized by an almost opposite average negative and positive sentiment, respectively. It is interesting to notice that the only average positive sentiment is associated to a topic shared by EXP and USTD, i.e. electrostimulation describing the same type of physical stimulus, but surrounded by a very different narrative (experimental, innovative VNS, versus traditional electroacupuncture).

**Table 4.** Sentiment by topic.

| Topic | max_sentiment | mean_sentiment | min_sentiment |
| --- | --- | --- | --- |
| 3 | 0.13 | -0.17 | -0.47 |
| 1 | 0.21 | -0.1 | -0.41 |
| 7 | 0.13 | -0.09 | -0.31 |
| 4 | 0.15 | -0.05 | -0.25 |
| 2 | 0.24 | -0.04 | -0.32 |
| 8 | 0.28 | -0.04 | -0.36 |
| 5 | 0.15 | -0.03 | -0.21 |
| 9 | 0.17 | -0.03 | -0.23 |
| 6 | 0.15 | -0.01 | -0.17 |
| 10 | 0.26 | 0.02 | -0.22 |

### 3.3. Emotions Analysis

When comparing the whole range of normalized emotions (Table 5), the three categories present two shared and dominant emotions, i.e. neutrality and approval, probably unsurprisingly for a scientific document.

**Table 5.**
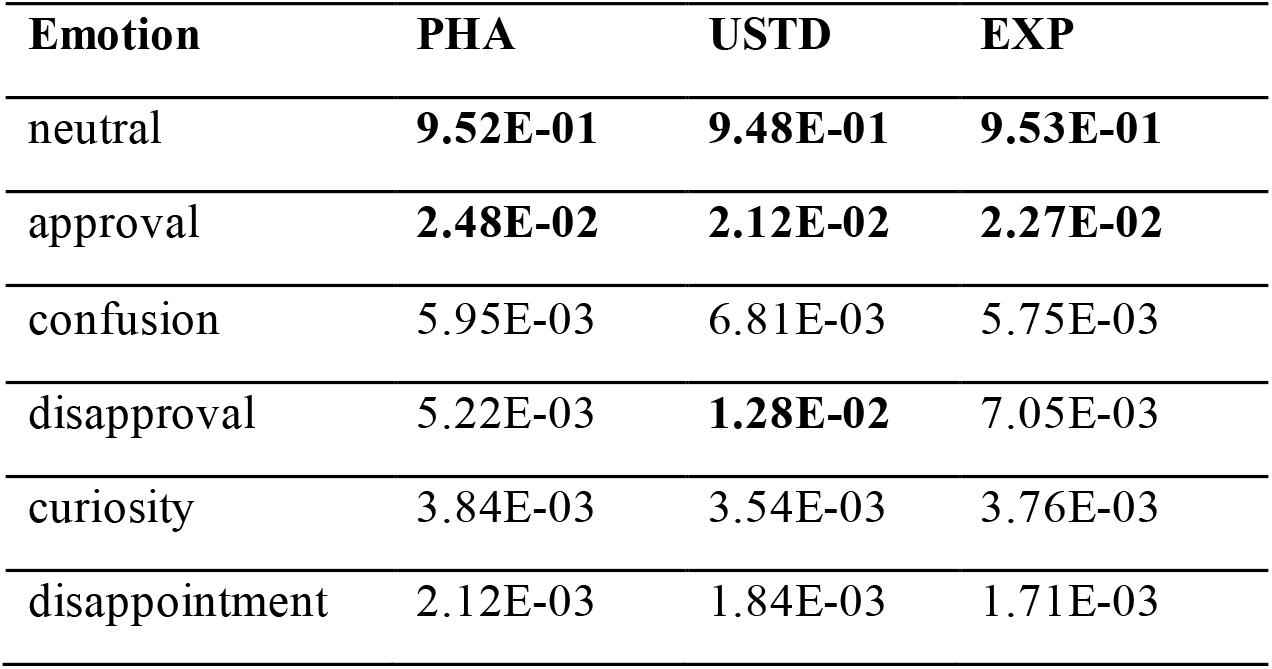
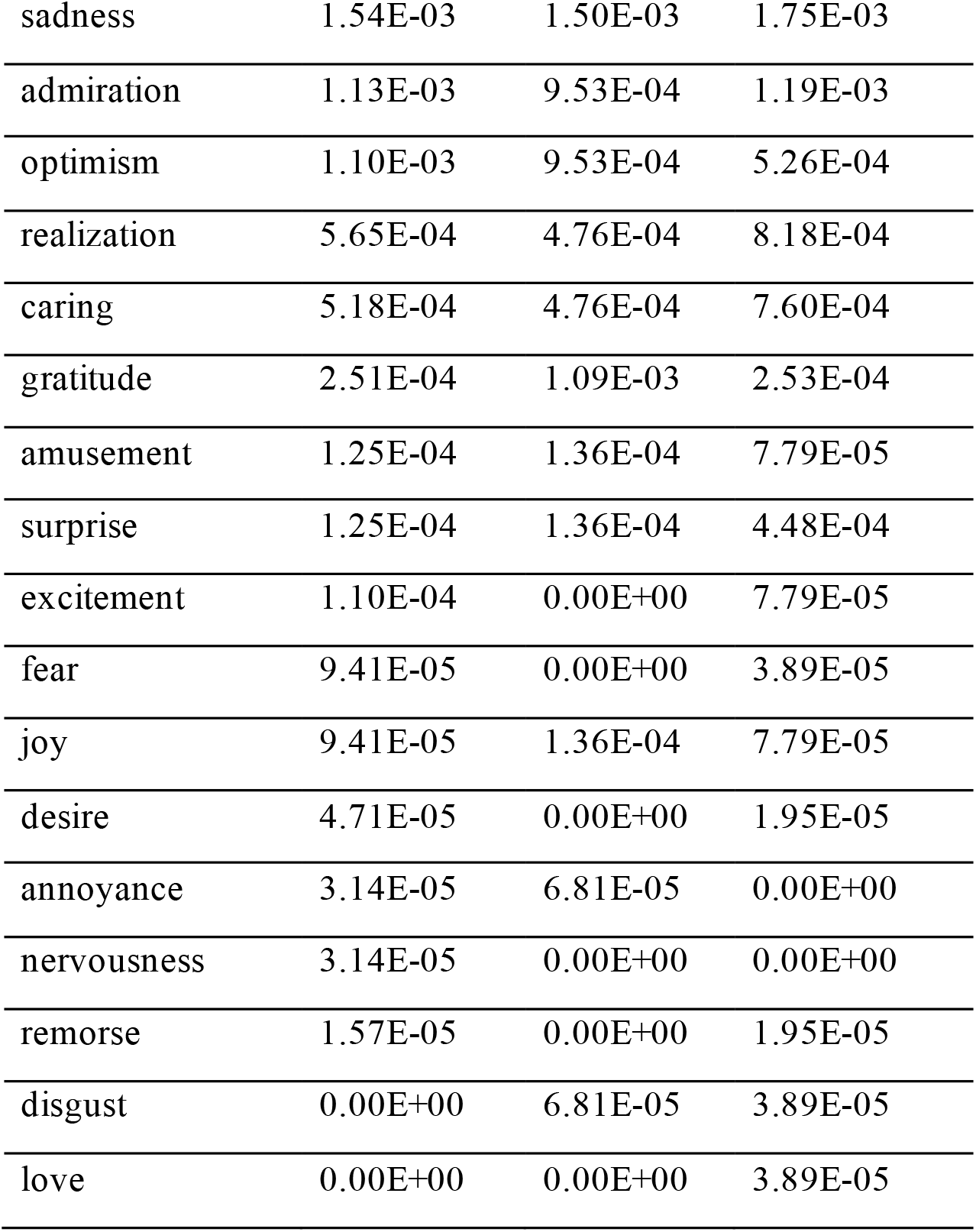
Ranked normalized emotions, in bold top ranking emotions, by category.

When looking for differences, few key observations arise: first, USTD presents with a smaller range of emotions, while EXP and PHA share the same variability in range, with however two complementary emotions: PHA lacks love and disgust, two strong emotions, in line possibly with a more neutral writing, and EXP lacks nervousness and annoyance possibly in agreement with the excitement for novelty. In USTD the lack of breadth in the emotional range seems compensated by a 3rd relatively relevant emotion: disapproval, which together with neutral and approval describe the emotions characterizing USTD writing.

When looking at the trend over the whole article (Figure 3) we observe that the seemingly uniformity of emotions presents indeed differential trends. In particular, while PHA and EXP confirm a similar emotional content, USTD presents a pattern that is fairly unique.

**Figure 3.**
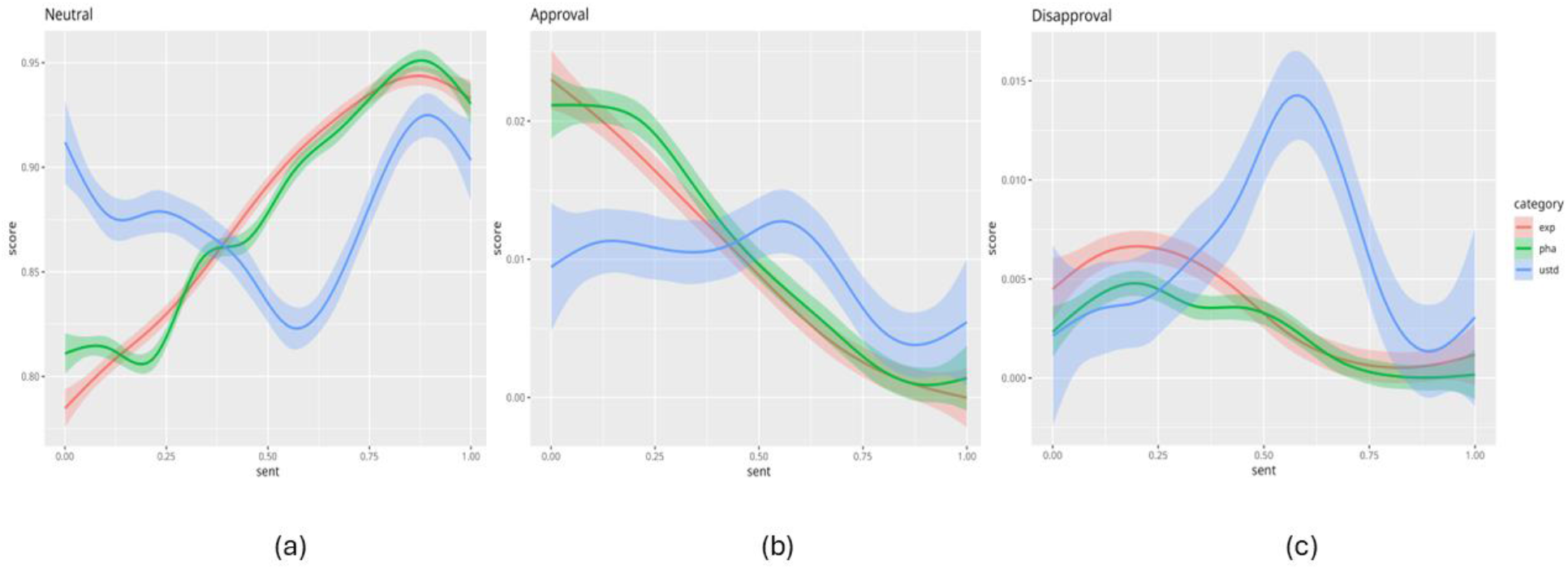
Dominant emotions: neutral, Approval and Disapproval in panels a-c respectively.

Neutrality appears to build and grow over the course of the writing in both PHA and EXP, while USTD loses neutrality over the first half of the discussion. Further, the lack of neutrality in PHA and EXP is compensated by approval to set the tone at the beginning of the reports, while the decrease in neutrality is compensated in USTD by disapproval. In other words, PHA and EXP build their scientific report by setting the tone with approval first, and move then to neutrality, while USTD starts with neutrality that quickly loses ground for approval/disapproval to go back to neutrality. It has to be noted that disapproval dominates over approval.

Although very different in substance, we can briefly discuss how these results complement the ones obtained in [10], in particular for the systematic observations on the three PHA, EXP and USTD categories. To ease the comparison, we report here (Figure 4) the results obtained for the systemic analysis that represents the 3 categories under study in terms of 6 so-called socio-anthropological variables. In this representation EXP and USTD appear to be more similar than EXP and PHA, and in particular 3 out of 6 variables have a shared value and in particular the spatialization of the disease, both in terms of the etiology and of the therapy (i.e. where the origin and the treatment of RA are supposed to be in term of body geography). This similarity does not emerge from the emotions analysis, since only for optimism can we observe the same (stable) trend for EXP and USTD (dropping in PHA), however this emotion contributes negligibly to the overall analysis (see all plots in Figure S1).

**Figure 4.**
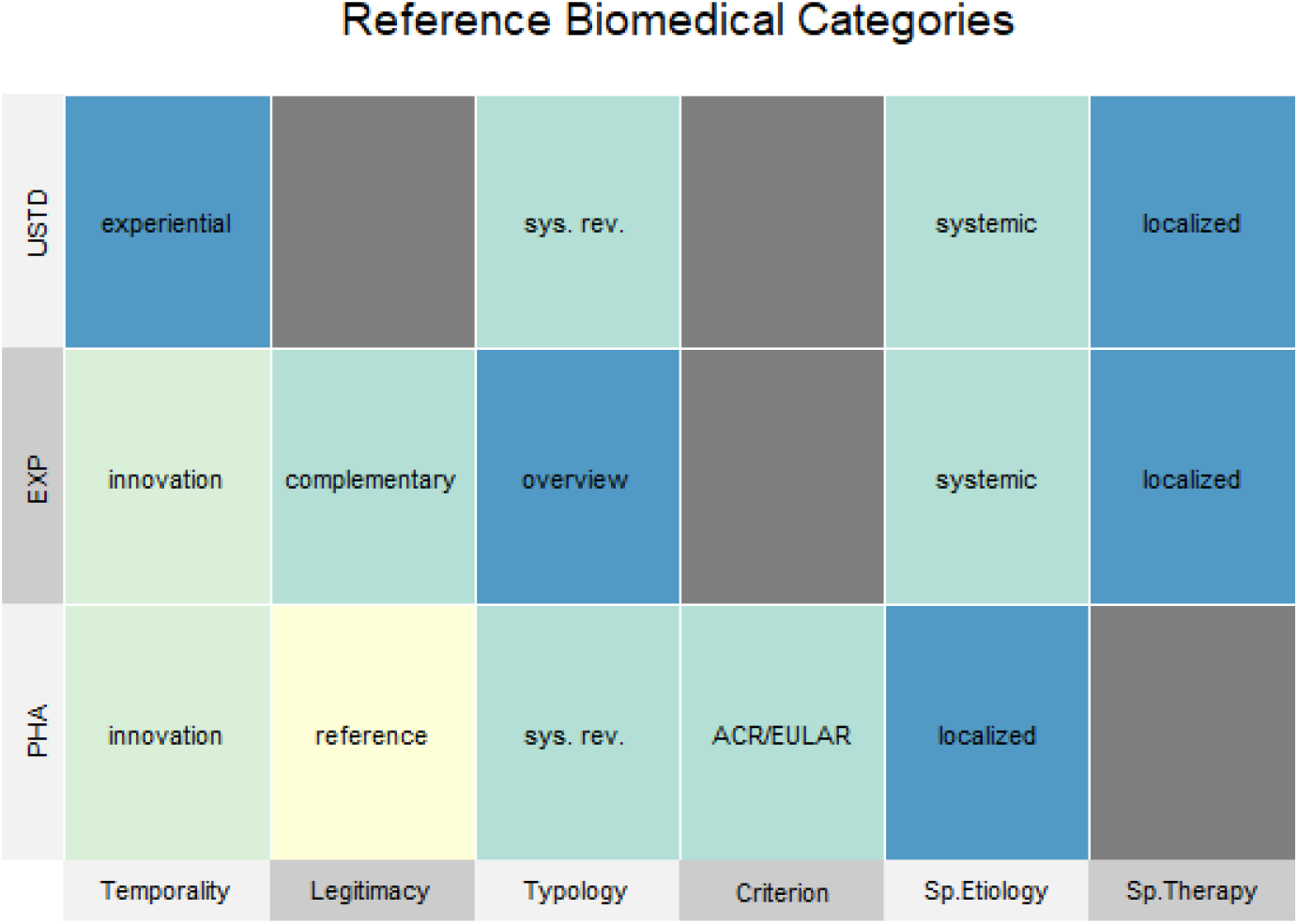
reproduced with permission from PLoS ONE Nardini et al. 2022. Representation of the 3 categories in terms of 6 socio-anthropological variables.

Conversely, PHA and EXP share the same approach on temporality (indicating how the proposed therapy relates to time, i.e. whether its solidity descend from long dated experience of from cutting-edge discoveries) which is innovative, versus the experiential for USTD, and we can speculate that this is the aspect captured by emotions. We could argue that supporting a medical approach based on cutting edge scientific discoveries (a requirement of modern medicine) legitimates the authors to drop neutrality (which is another requirement of scientific writing) in favor of approval to assertively remark the legitimacy of the discussion that follows. Conversely USTD, that relies on experience as a guarantee for legitimacy, is forced to back up this weakness with neutrality, at least to set the tone, i.e. at the beginning of the article. In summary, while PHA and EXP, strong of the scientific innovation they rely on, can drop one of the commitment/hallmarks of scientific writing (neutrality) USTD must make up for this original sin in opening by neutrality.

## 6. Conclusions

This analysis represents an automatic approach to the medical discourse and complements the manually curated work presented in [10].

Among the advantages, we observe that topic modeling was able to reproduce the majority of the sub-categories that were decided a priori, provided a sufficient number of articles exists. Based on this availability, indeed, the automatic approach clearly distinguished conventional and biologic drugs (all merged in the category AntiInflammatory). Intriguingly electrostimulation was identified in one topic only (Topic 10) including both electroacupuncture and VNS, that belong to two different categories (USTD and PHA, respectively), indicating that the two approaches share an amount of commonalities that automatic approaches can identify, despite limited to no communication among the two research areas. Finally, the emotions analysis was able to discover a similarity between PHA and EXP and versus USTD that we could relate to the temporal legitimation of the approaches, suggesting that the medical discourse is strongly biased by these variables.

On a less positive note, we encountered a significant challenge in needing/lacking a unified source for downloading scientific articles using automated scripts/services/API. Other efforts have already assessed the potential benefit and lost opportunities descending from enabling such sourcing [23,24].

In our specific case, this resulted in a time-consuming and labor-intensive process of managing and developing scripts to download articles from various sources using the paywall access from our institute. Owing to the incomplete openness of articles, we could fully download only 204 articles (58.8 % of the selected PMID). The Open Science movement should consider this, as not only human accessibility but also machine accessibility represents an obstacle. Indeed, the article’s various PDF/HTML formats need accurate methods to transform it into simple, machine-processable text. At the source, the articles are in WYSIWYG Microsoft Word-like, Latex, or Markdown formats that are more easily and precisely processable.

Openness in the source code of articles would allow researchers to quickly and easily extract information from large numbers of scientific articles. By automating the process of downloading source code articles, researchers can focus on higher-level tasks, such as data analysis and interpretation, rather than spending valuable time and effort downloading and cleaning the data.

This study showcases the potential of automated semantic analysis within RA research.

Overcoming the inherent data collection hurdles, we analyze a sizable corpus exceeding 200 articles. The findings illuminate the synergistic relationship between meticulous manual curation [10] and computer-assisted semantic analysis. This investigation serves as a springboard for future advancements. For instance, it posits the development of a novel framework that leverages the strengths of manual curation by transforming the curated data into a training set for a deep learning model. This framework has the potential to significantly extend the reach of manual curation beyond the constraints of human resources, enabling the analysis of considerably larger datasets in subsequent studies.

## Data Availability

All data produced in the present work are contained in the manuscript

## Supplementary Materials

The following supporting information can be found at www.sciepublish.com/xxx/s1, Fig. S1: Emotion analysis for all 27+1 emotions; Table S1: Table containing queries and full list of 500 articles originally returned; Table S2: List of 28 articles manually processed in the socio-anthropological analysis; Table S3: list of the 204 PMID processed in the automatic analysis; Table S4: Full list of the analyzed emotions; Table S5: Semanticase Topic Words.

## Author Contributions

Conceptualization, M.S. and C.N.; Methodology, M.S. and C.N.; Software, M.S.; Validation M.S. and C.N.; Formal Analysis, M.S. and C.N.; Data Curation, M.S. and C.N.; Writing – Review & Editing, M.S. and C.N.

## Notes

### Competing Interest Statement

The authors have declared no competing interest.

### Funding Statement

This study did not receive any funding

