## Supplementary material for "Large-scale analysis of the medical discourse on rheumatoid arthritis: complementing a socio-anthropologic analysis": Supplementary.docx

S1 Semanticase

1. Introduction

This supplementary section introduces the algorithmic core of Semanticase, developed by PiazzaCopernico srl, Semanticase represents a significant advancement in the field of natural language processing and semantic analysis for people with not a programming skills.. As a product of PiazzaCopernico srl (http://www.semanticase.it), this innovative web-based software suite is specifically designed to facilitate in-depth semantic exploration of textual data, catering to the diverse needs of scientific researchers and analysts across various disciplines. By providing an intuitive and user-friendly web interface, Semanticase seamlessly integrates a robust and versatile suite of algorithmic functionalities, thereby empowering users to uncover the complex semantic patterns and relationships underlying their textual data with unprecedented ease and precision. This supplementary section delves into the technical core of Semanticase, elucidating its algorithmic foundations and highlighting its capabilities for facilitating comprehensive semantic analysis.

The core analytical engine of Semanticase is built upon a customized implementation of the Structural Topic Model (STM). The STM transcends the limitations of traditional topic modeling techniques by identifying thematic clusters within textual data and uncovering the underlying hierarchical structure of these topics. This unveils the intricate relationships and interplay between thematic elements, fostering a more nuanced understanding of the semantic landscape within the corpus. Furthermore, the customized STM employed by Semanticase may incorporate specific modifications or optimizations meticulously tailored to enhance its performance on the types of textual data the software is designed to analyze.

By harnessing the power of this advanced topic modeling technique, Semanticase empowers researchers to delineate the dominant themes resident within their corpus and elucidate the intricate interplay and relationships between them. This facilitates a significantly more comprehensive exploration of the semantic richness embedded within the textual data under investigation.

2. Corpus Import and Manipulation

Semanticase fosters a streamlined workflow for corpus integration through its robust and versatile import and manipulation tools suite. This section details the functionalities that empower researchers to incorporate textual data from various sources into their analysis seamlessly.

2.1 Broad File Format Support and Specialized Content Ingestion

Semanticase leverages the Apache Tika library for comprehensive file format recognition. This enables the extraction of textual content from standard document formats (DOCX, PPTX, PDF), email archives, and web pages.

Furthermore, Tesseract OCR technology empowers the inclusion of valuable insights from scanned documents (images) or PDFs containing scanned content, expanding the scope of analyzable data to include physical documents or historical archives.

Beyond conventional file formats, Semanticase caters to the specific needs of diverse content types:

Customized import routines, built upon the 'officer' R package, meticulously handle complex elements like word art within presentations and documents for seamless PPTX and DOCX integration.

The advanced speech-to-text capabilities of wav2vec2-large are harnessed to transform audio files into usable textual data. This facilitates the incorporation of interviews, lectures, or other audio recordings into the analysis, enriching the corpus with valuable spoken content.

Additionally, there are routines to consider in the corpus covariates data, such as the time length between words and words’ speed distribution, which are helpful for further analysis.

Informal media data can be effortlessly integrated through a dedicated import module for WhatsApp chat archives, facilitated by the 'rwhatsapp' R package. This streamlines incorporating social interactions and communication patterns from WhatsApp conversations into the analysis.

2.2 Post-Import Quality Control and Corpus Translation

Semanticase prioritizes data quality and integrity through various features following the import process.

A dictionary-based approach meticulously identifies and addresses potential inconsistencies or errors within the imported content, ensuring the accuracy of the corpus, especially in OCR cases.

Semanticase Python and R built-from-scratch tools can cleanse information strings from residual text formatting or specific file format commands introduced during import. This standardization process optimizes the corpus data for subsequent analysis.

Recognizing the prevalence of multilingual research data, Semanticase incorporates functionalities for language detection and translation.

The platform leverages Google's Compact Language Detector 3 library to identify the language for each entry in the corpus, automatically putting it as an additional covariate in the corpus. Then Semanticase integrates the MarianMT transformer translation engine. This empowers researchers to seamlessly translate their corpus data into a desired language, dissolving language barriers and enabling the analysis of textual content from various sources.

By providing comprehensive import functionalities, meticulous quality control mechanisms, and multilingual support, Semanticase equips researchers with the tools to effectively integrate, prepare, and analyze textual data from diverse sources, fostering a robust foundation for in-depth semantic exploration.

2.3 Granular Text Exploration through Sentence-Level Processing

Semanticase extends its text manipulation capabilities to facilitate in-depth exploration at the sentence level. This functionality empowers researchers to dissect imported documents into individual sentences. Granular control over sentence units offers distinct advantages for specific analytical tasks. For instance, sentiment analysis often hinges on the sentiment expressed within individual sentences, where subtle variations can hold significant meaning. Semanticase leverages the established pyBSD library in the Python environment to achieve this segmentation of sentences. The pyBSD library provides a robust and efficient toolkit for text manipulation, ensuring accurate sentence-level processing within the Semanticase platform. By enabling sentence-by-sentence control over the corpus, Semanticase equips researchers with a powerful tool to delve deeper into the semantic nuances of their textual data, fostering an insightful analysis.

2.4 Statistics on Corpus

A thorough comprehension of the corpus's composition and characteristics forms the cornerstone of a successful semantic analysis.

Semanticase leverages the 'quanteda_textstat' R package to generate a rich tapestry of descriptive statistics on the corpus. These encompass fundamental metrics such as total word count, average document length, and the number of documents within the corpus. These foundational measures establish a baseline understanding of the corpus size and structure.

Beyond rudimentary descriptive statistics, Semanticase delves deeper into textual complexity by implementing advanced metrics calculated using the 'quanteda_textstat' package. A prominent example is the Simpson D index, a well-established and robust indicator of lexical diversity. This metric assesses the corpus's vocabulary richness, revealing the extent to which unique words are employed. The Simpson D index is generally considered adequate across a wide range of corpora, making it a valuable tool for gauging overall lexical variation.

Furthermore, Semanticase incorporates a readability measure derived from the Strain Index, specifically adapted for the analysis of textual data. The original Strain Index was designed to evaluate readability within texts of three sentences. Quanteda.textstats addresses this limitation by employing a normalized version of the Strain Index, enabling its application to corpora encompassing a broader spectrum of document lengths.

In addition to these core metrics, Semanticase generates a granular breakdown of the corpus composition. This includes statistics such as the total number of characters, sentences, tokens (individual words or meaningful units), unique tokens (excluding duplicates), punctuation marks, numeric tokens, symbols, tags, and emojis. This comprehensive overview empowers researchers to understand their corpus's specific elements, facilitating informed decisions during subsequent analysis stages.

It is imperative to acknowledge that the computational intensity of these statistical calculations scales with larger corpora. To ensure efficient processing and platform stability, Semanticase has implemented an empirical threshold of 2 GB for imported corpus size within the R environment. Corpora exceeding this size may encounter extended processing times. Alternative statistical analysis tools outside of Semanticase may be necessary for extensive datasets.

3. Semanticase Run’s Concept: Preserving the Analytical Journey

Within the domain of computational text analysis, the pathway from raw data to meaningful insights is often multifaceted, encompassing intricate data manipulation and analytical procedures. Semanticase champions a comprehensive approach to analysis, meticulously capturing each step of this transformative process. A cornerstone concept within Semanticase is the designation of a *run***.** A run transcends a mere analysis; it meticulously chronicles the entire analytical flow, encompassing, after the corpus import, all further data cleaning, manipulation, and analysis steps undertaken on a specific corpus. Each run serves as a unique and immutable record, faithfully preserving the sequence of transformations and parameter selections applied to the data.

This commitment to meticulous record-keeping grants researchers many advantages:

- *Unwavering reproducibility*: by meticulously documenting the entire analytical workflow within a designated run, Semanticase fosters the cornerstone principle of scientific inquiry-reproducibility. This empowers researchers to effortlessly replicate the analysis at a later date or transparently share the exact methodology with colleagues. This fosters trust and verification within the broader scientific community, solidifying the foundation of research findings.
- *Knowledge Preservation:* the detailed record of each run transcends a simple log; it transforms into a valuable repository of knowledge. Researchers can revisit past runs to reconstruct the analysis and glean insights into the rationale behind specific data manipulation or analysis parameter selections. This fosters informed decision-making during future analyses and streamlines refining analytical approaches.
- *Empowering Human-in-the-Loop Refinement*: the ability to revisit and meticulously analyze past runs allows researchers to adopt a human-in-the-loop approach to analysis refinement. By critically examining the outcomes of previous runs, researchers can pinpoint areas for improvement and iteratively refine their analytical approach. This cyclical process leads to the development of increasingly nuanced and insightful results, propelling research endeavors toward greater robustness and reliability.

Semanticase's runs give a rigorous and well-documented analytical environment. This meticulous approach promotes transparency, encourages knowledge retention, and empowers researchers to continuously refine their analyses, ultimately leading to more robust and impactful research.

In addition, to improve the readability of models’ results, Semanticase, in every run implementation, considers not only singular but also all possible N-grams (N=1,2,3).

In Semanticase, N-grams represent contiguous sequences of N words extracted from a text sample.

3.1. Synonyms and Stopwords: Refining the Corpus Vocabulary

Tailoring the corpus vocabulary is pivotal in optimizing subsequent semantic analysis within Semanticase. This section explores the functionalities designed to refine the corpus vocabulary by managing stopwords and synonyms.

*Stopwords* encompass frequently occurring words with minimal semantic value in the context of analysis. Examples include common prepositions (e.g., "the", "a") and conjunctions (e.g., "and", "but") or month, day names and abbreviations, and numbers. While essential for grammatical structure, these words often contribute limited meaning to the overall thematic content. Other stopwords can be largely diffused, already known and identifying the corpus. As a pure example, *wikipedia* word can be a stopword in an analysis of Wikipedia entries. Semanticase empowers researchers to selectively eliminate stopwords based on the specific goals of their study. This streamlining process reduces noise within the corpus and fosters a more focused exploration of the substantive semantic content.

Beyond stopwords, *synonyms* – words that share similar meanings in the analysis objective– can present a distinct challenge for semantic analysis. If left unaddressed, the presence of synonyms can lead to the artificial inflation of specific themes within the corpus. To address this, Semanticase offers functionalities for synonym management. Researchers can leverage the built-in resource editor to construct custom synonym lists meticulously. This empowers them to consolidate synonymous terms under a single representative term, fostering a more accurate representation of the underlying thematic structure within the corpus.

Semanticase employs regular expressions to execute synonym substitution and stopword elimination efficiently. Regular expressions offer a powerful and versatile tool for pattern matching within text, enabling the precise identification and manipulation of targeted words or phrases. Furthermore, Semanticase incorporates CPU parallelism to expedite these operations, ensuring efficient processing even for large corpora.

The management of stopwords and synonyms occurs within the designated run definition. This meticulous record-keeping approach ensures transparency and enables researchers to revisit and refine these choices during the human-in-the-loop analysis refinement process. Researchers can iteratively optimize their approach by critically evaluating the impact of stopword removal and synonym consolidation on the analysis outcomes. This leads to a more nuanced and insightful exploration of the corpus's semantic landscape.

The creation of the run gives the possibility to store the original text as uploaded to Semanticase (raw_data), keep the imported text (corpus) safe, and then store the modified text with stopwords and synonyms (run).

3.2. Run Statistics

Following the execution of an analysis run within Semanticase, a comprehensive suite of run statistics is calculated. Mirroring the corpus statistics functionality, run statistics provide valuable insights specific to the current analytical workflow. Like corpus statistics, run statistics adhere to an empirical threshold of 2 GB for R object size to ensure efficient processing within the platform.

A prominent component of run statistics involves the distribution of N-gram (N=2,3) frequencies. These co-occurrence patterns, also called *collocations*, can reveal thematic relationships and frequently used phrases within the text data.

Furthermore, run statistics encompass the *entropy* of both documents and individual words. In the context of documents, higher entropy indicates greater variability in word usage, suggesting a richer and more diverse vocabulary. Conversely, lower document entropy signifies a more limited vocabulary and potentially repetitive content. Similarly, word entropy assesses the level of predictability associated with individual words within the corpus.

4 Statistical Analysis

Here, we present the core of Semanticase statistical analysis of datasets with text.

Beyond the intrinsic thematic structure and word usage patterns within a corpus, a comprehensive understanding of textual data necessitates exploring external factors' influence. These external factors, termed *covariates*, encompass measurable characteristics associated with documents within the corpus. Covariates can be numerical (e.g., publication year) or categorical (e.g., author demographic, document genre). By incorporating covariates into the statistical analysis framework, researchers can illuminate potential relationships between these external characteristics and the content of the analyzed texts. This section delves into the robust suite of statistical analyses offered by Semanticase, specifically designed to elucidate the interplay between textual content and various covariate values. These analyses empower researchers to identify statistically significant differences between texts associated with distinct covariate values, fostering a more nuanced understanding of the factors shaping the observed semantic landscape.

4.1. Word Frequency and Keyness Analysis

This subsection delves into the preliminary exploratory analysis to elucidate the interplay between word usage and covariates. Semanticase analyzes covariates with a minimum of two distinct values and a maximum of ten categories to ensure computational efficiency and interpretable results. This range strikes a balance between flexibility and analytical tractability.

As a foundational step, Semanticase facilitates the identification of the most frequent words associated with each covariate value. This initial analysis empowers researchers to gain a rapid grasp of the vocabulary distribution across different covariate categories. The system furnishes a list of the most frequently occurring words for each distinct covariate value. This information empowers researchers to discern potential thematic variations or vocabulary biases associated with specific covariate categories.

Complementing the initial investigation into frequent words per covariate value, Semanticase incorporates *keyness* analysis to provide researchers with a more nuanced understanding of word usage patterns. The keyness metric is operationalized within the textstat_keyness function from the quanteda.textstat R package, quantifying the relative frequency of a word within a specific corpus segment relative to its frequency in a designated reference corpus. Within the context of covariate analysis, Semanticase adopts a targeted approach. The target group encompasses documents associated with a single covariate value, whereas the reference group comprises the combined word frequencies across all other documents in the corpus. This strategic selection aims to identify words exhibiting statistically significant higher frequency within a particular covariate category compared to the overall corpus baseline. Such words, pinpointed through keyness analysis, potentially reflect that specific covariate category's thematic focus or distinctive vocabulary characteristic.

The keyness values for each word are a chi-squared statistic calculated within the textstat_keyness function. Furthermore, Yates' correction is applied to this chi-squared statistic to mitigate potential biases in situations with limited sample sizes within individual documents. By integrating keyness analysis with covariate values, Semanticase empowers researchers to identify statistically more prominent words within specific document subsets defined by covariate categories. For a more comprehensive exploration of keyness analysis and its applications in text analysis, we refer interested readers to [17].

4.2. WordNet Analysis

Semanticase expands its covariate analysis repertoire by incorporating WordNet, a well-established lexical database of semantic relationships. Semanticase leverages the *textnets* library within the Python environment for each distinct covariate value to construct a dedicated bipartite graph. These bipartite graphs visually represent the semantic relationships between content words (nouns, verbs, adjectives, adverbs) extracted from documents associated with that specific covariate value and their corresponding synsets (sets of synonyms) within WordNet.

By critically examining these covariate-specific bipartite graphs, researchers gain valuable insights into the semantic organization of the corpus vocabulary. Prevalent synsets within a graph highlight dominant thematic elements characterizing the corresponding covariate category. Thematic clusters, visualized through interconnected nodes, expose areas of particular emphasis within the covariate-specific vocabulary. .

4.3. Sentiment and Emotion Analysis

Semanticase incorporates functionalities for sentiment and emotion analysis. This multifaceted approach empowers researchers to go into the affective dimensions (emotions and opinions) embedded within the corpus.

The initial cornerstone of Semanticase's sentiment analysis leverages a lexicon-based approach implemented within the *sentimentr* R package. This method relies on pre-defined sentiment lexicons, encompassing words and phrases categorized as positive, negative, or neutral. Semanticase furnishes researchers with an overall sentiment polarity for the corresponding document subset by analyzing the frequency of these sentiment-laden elements within each covariate category. This analysis provides valuable insights into the general emotional tone and prevailing opinions associated with specific covariate values. Sentiment interpretation is inherently nuanced, and *valence shifters* (e.g., negation words, intensifiers, adversative conjunctions) can significantly alter the intended sentiment. While *sentimentr* incorporates rules to mitigate the influence of some valence shifters, researchers should exercise caution when interpreting the results, acknowledging these potential limitations.

Complementing the lexicon-based sentiment analysis, Semanticase offers a machine-learning approach for sentiment category classification. This functionality utilizes the *nlptown/bert-base-multilingual-uncased-sentiment* model within the HuggingFace pipeline in Python environment. This pre-trained model allows researchers to classify sentiment on a 1-to-5 scale, with 1 signifying negative sentiment and 5 signifying positive sentiment. This fine-grained rating system offers a more nuanced understanding of sentiment variations within the corpus, potentially revealing subtle differences in opinion across covariate categories.

Expanding beyond sentiment analysis, Semanticase incorporates functionalities for emotion classification. This analysis leverages the *joeddav/xlm-roberta-large-xnli* model within the HuggingFace pipeline for a zero-shot learning approach. Zero-shot learning empowers the model to classify emotions within the text data without explicit training on emotion-labeled examples. The model can identify five core emotions: *anger, disgust, fear, joy, and sadness*. Furthermore, it exhibits multilingual capabilities, adapting to the language of the text (English, Italian, Spanish, French, or German). By calculating the relative frequency of each identified emotion within documents associated with specific covariate values, Semanticase empowers researchers to discern potential emotional undercurrents associated with different covariate categories.

Using covariate one can quantitatively estimate the emerging thematic structures (a.k.a. *topics*) associated with different article types within the corpus. Incorporating this covariates gives more profound insights into how thematic structures vary across differen categories. In particular, STM used two defined statistics to quantify this insight:

5. Topic Modeling

Corpus analysis transcends the exploration of individual words and their co-occurrence patterns. Topic modeling aims to identify latent thematic structures residing within the textual data. Semanticase incorporates Structural Topic Modeling (STM), a powerful and versatile approach to uncovering these underlying thematic trends.

At its core, STM operates under the assumption that documents within a corpus can be represented as mixtures of a finite number of underlying topics. Each topic encompasses a thematic cluster characterized by a probability distribution over the words that frequently co-occur within that theme and a probability distribution of each topic between the documents. By analyzing this thematic structure, researchers gain valuable insights into the overarching themes and latent semantic relationships embedded within the corpus. Topic modeling empowers researchers to identify recurring thematic threads, classify documents based on their composition, and explore the thematic relationships between different document subsets.

5.1 Topic Content and Prevalence in Semanticase's STM

Semanticase's implementation of STM furnishes researchers with two fundamental insights regarding the identified topics: content and prevalence.

*Topic prevalence* refers to the degree to which a particular topic is represented within a document or across the entire corpus. It measures how frequently or prominently a topic appears in a text, and using STM, we measure topic by topic whether there is a stronger or weaker association with a specific covariate value.

*Topic content* encompasses the specific words that define a particular topic. It represents the vocabulary that characterizes a concept or a theme. STM defines the probability for each word in the topic and to be associated with each covariate. Importantly, this enables the possibility that few words in a topic are more (high probability) or less (low probability) associated with a specific category. This indicates that, although the topic is important across the whole corpus, i.e., shared among all covariate values, different words may be characteristic and descriptive of this topic, covariate-wise. I.e., a corpus topic can be described by a partially different vocabulary, covariate-wise.

5.2 Semanticase STM’s Customization

Semanticase's STM functionality incorporates several customized key features designed to enhance usability and analytical efficiency.

*Automatic Topic Number Estimation*: A crucial aspect of topic modeling involves determining the optimal number of topics (K) to capture the underlying thematic structure without overfitting the data. Semanticase addresses this challenge by employing Automatic K Number Estimation using Singular Value Decomposition (SVD). This automated approach streamlines the topic number selection process, empowering researchers to focus on interpreting the derived thematic landscape.

*N-gram Vocabulary*: Semanticase leverages an N-gram vocabulary (N=1,2,3) within the STM analysis. This approach incorporates not only individual words (unigrams) but also bigrams and trigrams. By including these N-grams, Semanticase captures thematic nuances reflected in the co-occurrence patterns of terms, potentially leading to a more refined representation of the underlying thematic structure.

*Enhanced Graph Visualization*: Semanticase offers interactive topic relationship visualizations rendered in HTML to facilitate the interpretation of topic modeling results. These visualizations, generated using the *ggplot* and *ggiraph* R packages, depict the relationships between topics, empowering researchers to explore the thematic coherence and potential thematic hierarchies within the corpus. Following the execution of the topic model, Semanticase exploits the STM inter-topic correlations (directly output from the model) as measures of similarity among the topics and constructs a hierarchical clustering tree. Using aggregative logic, each leaf is recursively associated with the most probable words that are shared between the topics’ group.

*Interpretation and Utilizing Topic Model Results*: By critically examining the topic content and prevalence information, researchers can assign labels and descriptions to each topic, fostering a meaningful understanding of the latent thematic structure within the corpus.

*Topic Sentiment*: Semanticase extends its analytical capabilities by integrating topic modeling with sentiment analysis. By leveraging mean document-level sentiment scores and the probability distribution of documents across topics, Semanticase calculates a weighted mean sentiment for each topic. This metric quantitatively indicates the prevailing emotional tone or sentiment that characterizes a particular topic. To ensure the reliability of these sentiment estimates, Semanticase incorporates confidence intervals around the weighted mean sentiment values.

Acknowledge: The creation of Semanticase will not be possible without the valuable effort of the R&D team of PiazzaCopernico srl: Daniela Pellegrini (from beginning), Marcello Pucci, Alessandro Dell'Orto (past), Lorenzo Pozzi and Sara Zuzzi.
